# Potential Efficacy of Streptomycin in Amikacin-resistant *Mycobacterium avium-intracellulare* Complex Pulmonary Disease

**DOI:** 10.64898/2026.04.03.26350100

**Authors:** Tatsuya Kodama, Kozo Morimoto, Yoshiro Murase, Akio Aono, Koji Furuuchi, Keiji Fujiwara, Masashi Ito, Takashi Ohe, Fumiya Watanabe, Kinuyo Chikamatsu, Shiomi Yoshida, Yusuke Minato, Yoshiaki Tanaka, Miyako Hiramatsu, Yuji Shiraishi, Takashi Yoshiyama, Satoshi Mitarai

## Abstract

Aminoglycoside drugs, amikacin, streptomycin, and amikacin liposome inhalation suspension are crucial for treating refractory *Mycobacterium avium-intracellulare* complex pulmonary disease. In *Mycobacterium tuberculosi*s, cross-resistance occurs between amikacin and kanamycin, but not between amikacin and streptomycin in genetic drug susceptibility testing. However, the occurrence of cross-resistance among aminoglycosides remains unclear in *M. avium-intracellulare* complex. We aimed to evaluate cross-resistance among aminoglycosides to determine whether streptomycin or kanamycin remains effective after the development of amikacin resistance. This single-center retrospective study included 20 patients with amikacin-resistant *M. avium-intracellulare* complex harboring *rrs* mutations. Paired analyses of streptomycin and kanamycin minimum inhibitory concentration values before and after amikacin resistance development were performed. In addition, streptomycin– and kanamycin-resistant strains were generated *in vitro* and resistance-associated mutations were identified using whole-genome sequencing. No significant increase was observed in streptomycin minimum inhibitory concentration values following amikacin resistance. In contrast, kanamycin values uniformly increased to >256 µg/mL after the acquisition of amikacin resistance. Furthermore, amikacin– and kanamycin-resistant isolates shared mutations at position 1408 in the *rrs* gene, whereas streptomycin-resistant isolates exhibited mutations at position 20 in the *rrs* gene. These results suggest that amikacin and kanamycin exhibit cross-resistance in *M. avium-intracellulare* complex, whereas amikacin and streptomycin may not. Two cases in our cohort in which streptomycin treatment was effective after the acquisition of amikacin resistance further support these findings. In conclusion, streptomycin may be a potential therapeutic alternative for amikacin-resistant *M. avium-intracellulare* complex pulmonary disease. Future studies correlating streptomycin minimum inhibitory concentration values with clinical outcomes are required.

## INTRODUCTION

Globally, the prevalence and incidence of nontuberculous mycobacterial pulmonary disease have been increasing. This increasing pattern is also evident in Japan (1–3). *Mycobacterium avium-intracellulare* complex (MAC) is the predominant causative pathogen responsible for approximately 90% of nontuberculous mycobacterial pulmonary disease cases in Japan (2).

For cavitary, advanced/severe bronchiectatic, or macrolide-resistant MAC pulmonary disease (MACPD), parent amikacin (AMK) or streptomycin (SM) are recommended as aminoglycoside (AMG) drugs which are added to the initial treatment (4). In cases of refractory MACPD, amikacin liposome inhalation suspension may be added to the treatment in addition to AMK and SM (4). In Japan, kanamycin (KM) was recommended for treating patients with refractory MACPD until the revised nontuberculous mycobacterial pulmonary disease statement was issued in 2023 (5).

According to the latest Clinical and Laboratory Standards Institute (CLSI) guidelines, AMK resistance and amikacin liposome inhalation suspension resistance are defined as AMK minimum inhibitory concentration (MIC) values of ≥ 64 μg/mL and AMK MIC ≥128 μg/mL, respectively, when measured using cation-adjusted Mueller–Hinton broth (CAMHB) (6). Recently, cases of AMK-resistant MACPD have been increasingly reported (7–12). Regarding genotypic AMK resistance, mutations in the *rrs* gene, including those at positions 1408 and 1491, have been identified in AMK resistant isolates (7–12).

Clinically, AMK-resistant MACPD is of particular concern owing to its poor prognosis, especially when it occurs in macrolide-resistant cases. A previous study reported a 3-year mortality rate of 33% after the confirmation of AMK resistance (12). This poor outcome may be explained by the finding that clarithromycin (CAM) resistance has been identified as a risk factor for the development of AMK resistance (7, 12), which further limits the therapeutic alternatives. Therefore, it is clinically important to determine whether other AMG drugs, particularly SM and KM, remain effective after confirming AMK resistance.

In *Mycobacterium tuberculosis,* genotypic drug susceptibility testing suggests that AMK and KM exhibit cross-resistance, whereas no cross-resistance has been observed between AMK and SM (13–19). Although a previous study on MAC demonstrated that two missense mutations, Lys88Arg in *rpsl* and Pro138Ala in *rsmG* are key genetic determinants of high SM MIC values (20), their clinical relevance remains unclear.

The primary objective of this study was to assess cross-resistance among AMG drugs in MAC. Specifically, we compared SM and KM MIC values in MAC strains before and after the acquisition of AMK resistance and analyzed resistance-associated genetic mutations for each AMG drug.

## MATERIALS AND METHODS

### Study design and Study population

This single-center study included patients with MACPD who acquired AMK resistance through *rrs* mutations. This study was conducted at Fukujuji Hospital, Japan Anti-Tuberculosis Association, a 340-bed facility located at the northwest of Tokyo, Japan, between October 2021 and March 2025. We retrospectively reviewed the medical records of eligible patients.

Comparison of the SM and KM MIC values before and after AMK resistance confirmation was performed using both CAMHB containing 5% OADC and Middlebrook 7H9 broth containing 10% OADC for cases in which isolates before and after the acquisition of AMK resistance were available or for cases with known historical data.

### Microbiological examination for AMK, SM and KM

Antimicrobial susceptibility testing of AMK, SM, and KM was performed according to the CLSI-recommended broth microdilution method. Transparent MAC colonies grown on Middlebrook 7H10 agar were inoculated into CAMHB and Middlebrook 7H9 broths. The inoculum was standardized to match the 0.5 McFarland standard using a nephelometer. MICs were determined after incubation at 35 °C room air for 7 days. The tested concentrations of AMK were in the range of 0.25–256 μg/mL, and MIC values were interpreted both considering and ignoring the trailing growth (21). The SM and KM MIC values were also measured using the same procedure.

### Generation of SM and KM-resistant strains by serial passage and drug susceptibility testing

*M. avium* strain 63A, isolated from a patient with MACPD prior to treatment, was used to select high-level MIC value of SM and KM by serial passage. A total of 10^6^ cfu/mL of 63A (OD: 0.05) was suspended in CAMHB containing 5% OADC. Once it has grown to OD 0.2 (equivalent to 10^8^ cfu/mL), the strain equivalent to 2×10^6^ cfu was cultured on a 7H10 agar plate containing 16μg/mL of SM and KM by streak culture. Once the colony grew, cultivation was performed for the colonies again to a 7H10 agar plate containing 16µg/mL of SM and KM by streak culture. After the growth was observed, the drug susceptibility testing for AMK and SM were determined using CAMHB containing 5% OADC and Middlebrook 7H9 broth.

### Whole-genome sequencing

A 0.2 mL culture suspension was subcultured on Ogawa medium (Kyokuto, Osaka, Japan), which was incubated for 4 weeks or until confluent MAC growth was observed. DNA was extracted from loopful colonies grown on solid media using the conventional phenol-chloroform extraction method after bead beating (0.2 mm glass beads; Vortex Mixer GENIE2 at maximum speed for 7 min) (22). DNA concentration was measured using a Qubit (Thermo Fisher Scientific).

A genomic library was constructed from the total geno DNA of each MAC isolate and then subjected to paired-end sequencing on a NextSeq 550 (Illumina, San Diego, CA) platform generating 150-bp read lengths and converted to “fastq” format at the Research Institute of Tuberculosis, Japan Anti-Tuberculosis Association (23).

### Ethics statement

This study was approved by the Ethics Committee of the Fukujuji Hospital (approval no. 25045). This retrospective observational study was reported in accordance with the STROBE guidelines. Written informed consent was not required because of the retrospective observational study. However, the study information was provided to eligible patients through the Internet, and they were given the opportunity to withdraw from the study.

### Statistical analyses

Shapiro–Wilk test was used to assess data distribution. Continuous variables are presented as medians (interquartile ranges) for non-normally distributed data. Potential cross-resistance between AMK, SM, and KM was evaluated by comparing SM and KM MIC values before and after the acquisition of AMK resistance. MIC values for SM and KM were log₂-transformed, and within-isolate changes (ΔSM and ΔKM) were analyzed using paired Wilcoxon signed-rank test. In addition, equivalence was assessed using the two one-sided test with a prespecified equivalence margin of ±1 log₂ step, consistent with the commonly accepted essential agreement criterion used in MIC-based susceptibility testing method comparisons (24). All analyses were performed using STATA (version 17.1; StataCorp LLC; College Station, TX, USA).

## RESULTS

### Patients and characteristics of the study population

A total of 20 patients with AMK-resistant MACPD with *rrs* mutations were included in this study. All 20 isolates exhibited an AMK MIC value >256 μg/mL. Of the 20 cases, 19 were caused by *M. avium* and one by *M. intracellulare*. Fourteen of the 20 patients (70%) exhibited CAM-resistance at the time of AMK resistance. Regarding history of AMG use, the most common group comprised patients who received only AMK (50%,10/20), followed by two patients (10%) who received only KM. None of the patients received SM alone (Table 1). The *rrs* gene mutations were distributed as follows: 18 isolates (90%) harbored a mutation at position 1408, one isolate (5%) harbored a mutation at position 1491, and one isolate (5%) harbored a mutation at both positions 1408 and 203.

**Table 1.**
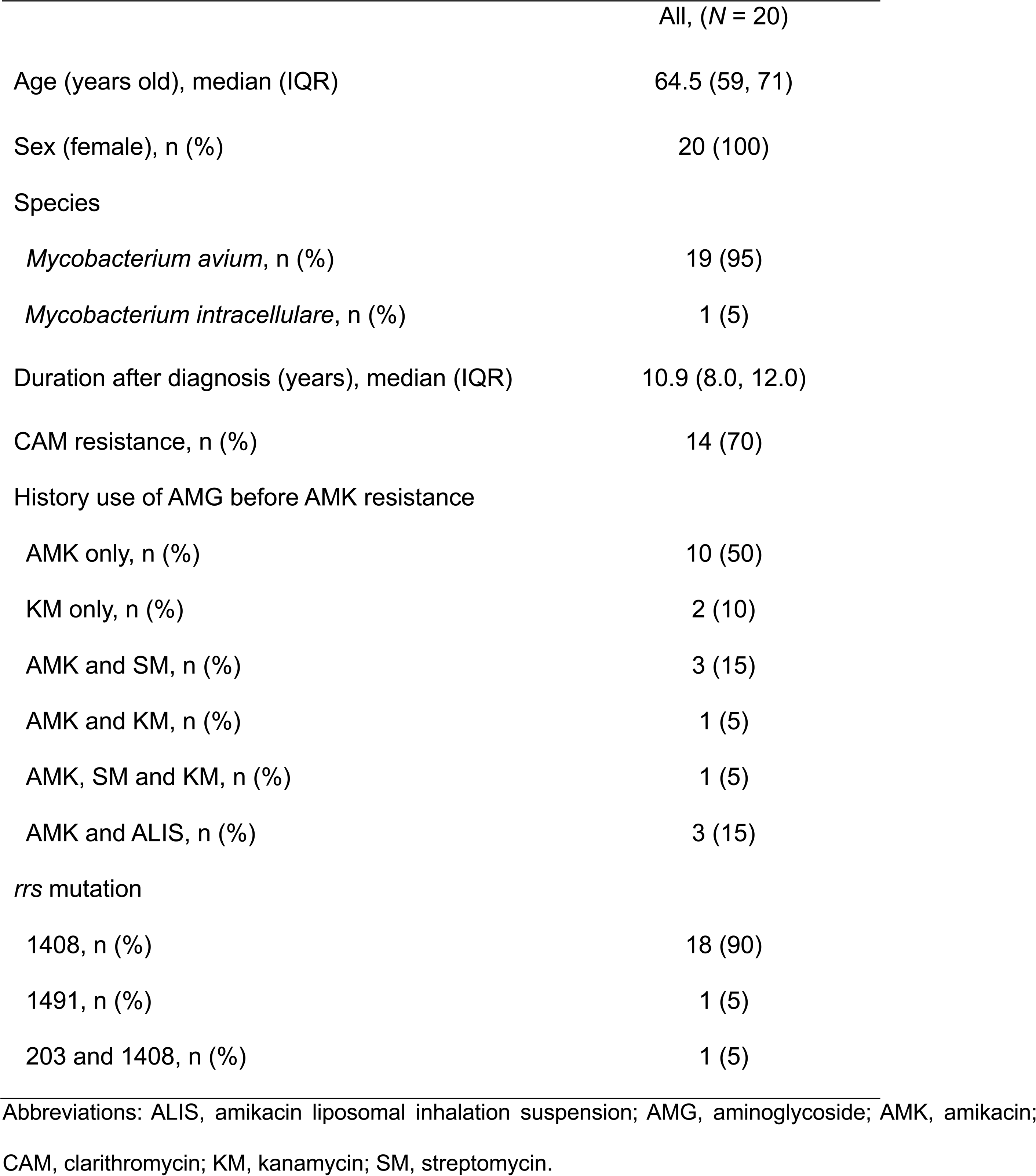
Characteristics of the study patients at AMK resistance confirmation.

### Comparison of the SM and KM MIC values before and after the confirmation of AMK resistance

Table 2 presents a paired comparison of the SM and KM MIC values measured in Middlebrook 7H9 broth (15 cases) and CAMHB (14 cases) before and after the acquisition of AMK resistance associated with *rrs* mutations, using either preserved isolates or data obtained from other hospitals.

**Table 2.** MIC results for AMK, SM, and KM in patients with AMK-resistant MACPD treated with 7H9 broth and CAMHB.

| Case | Species | MIC values (µg/mL) |  |  |  |  |  |  |  |  |  |  |  |
| --- | --- | --- | --- | --- | --- | --- | --- | --- | --- | --- | --- | --- | --- |
|  |  | 7H9 broth |  |  |  |  |  | CAMHB |  |  |  |  |  |
|  |  | Before resistance |  |  | After resistance |  |  | Before resistance |  |  | After resistance |  |  |
|  |  | AMK | SM | KM | AMK | SM | KM | AMK | SM | KM | AMK | SM | KM |
| 1 | *M. avium* | 8 | 16 | 8 | >256 | 16 | >256 | 8 | 8 | 4 | >256 | 4 | >256 |
| 2 | *M. avium* | 16 | 8 | 8 | >256 | 16 | >256 | 4 | 16 | 4 | >256 | 32 | >256 |
| 3 | *M. avium* | 4 | 8 | 4 | >256 | 4 | >256 | 8 | 8 | 8 | >256 | 16 | >256 |
| 4 | *M. avium* | 8 | 16 | 4 | >256 | 8 | >256 | 16 | 32 | 16 | >256 | 16 | >256 |
| 5 | *M. avium* | 16 | 4 | 8 | >256 | 2 | >256 | 32 | 16 | 32 | >256 | 4 | >256 |
| 6 | *M. avium* | 4 | 8 | 4 | >256 | 8 | >256 | 16 | 32 | 8 | >256 | 16 | >256 |
| 7 | *M. avium* | 8 | 8 | 8 | >256 | 8 | >256 | 32 | 16 | 16 | >256 | 32 | >256 |
| 8 | *M. avium* | 8 | 2 | 4 | >256 | 4 | >256 | 8 | 16 | 8 | >256 | 16 | >256 |
| 9 | *M. avium* | 16 | 32 | 8 | >256 | 32 | >256 | 8 | 32 | 4 | >256 | 32 | >256 |
| 10 | *M. avium* | 8 | 16 | 8 | >256 | 32 | >256 | 8 | 8 | 8 | >256 | 16 | >256 |
| 11 | *M. avium* | 32 | 16 | 32 | >256 | 8 | >256 | 4 | 8 | 2 | >256 | 8 | >256 |
| 12 | *M. avium* | 4 | 4 | 2 | >256 | 4 | >256 | 4 | 4 | 4 | >256 | 8 | >256 |
| 13 | *M. avium* | 4 | 2 | 4 | >256 | 16 | >256 | 8 | 4 | 4 | >256 | 8 | >256 |
| 14 | *M. avium* | 2 | 8 | 2 | >256 | 4 | >256 | 4 | 8 | 4 | >256 | 8 | >256 |
| 15 | *M. avium* | 2 | 4 | 2 | >256 | 4 | >256 | 32 | 4 | 32 | >256 | 4 | >256 |
| 16 | *M. avium* | 32 | 8 | 32 | >256 | 4 | >256 |  | No data |  | >256 | 4 | >256 |
| 17 | *M. avium* |  | No data |  | >256 | 16 | >256 |  | No data |  | >256 | 4 | >256 |
| 18 | *M. avium* |  | No data |  | >256 | 8 | >256 |  | No data |  | >256 | 32 | >256 |
| 19 | *M. avium* |  | No data |  | >256 | 8 | >256 |  | No data |  | >256 | 16 | >256 |
| 20 | *M. intracellulare* | 2 | 4 | 4 | >256 | 4 | >256 | 8 | 16 | 4 | >256 | 16 | >256 |

Abbreviations: MIC, minimum inhibitory concentration; AMK, amikacin; CAMHB, cation-adjusted Mueller-Hinton broth; KM, kanamycin; MACPD,
*Mycobacterium avium-intracellulare* complex pulmonary disease; SM, streptomycin

Regarding SM MIC values, both Middlebrook 7H9 broth and CAMHB showed highly consistent results. In 7H9, the changes in SM MIC values (ΔSM = log₂MIC_after – log₂MIC_before) were symmetrically distributed around 0, with a mean difference of 0 log₂ (SD = 1.01). The Wilcoxon signed-rank test indicated no significant shift in SM MIC values (p = 0.63), and the 90% CI of the mean difference (−0.55–0.55) fell within the predefined equivalence margin (±1 log₂ step). Similarly, in CAMHB, the ΔSM values were centered around 0, with a mean difference of +0.06 log₂ (SD = 0.93). No significant change was detected by the Wilcoxon signed-rank test (p = 0.64), and the 90% CI of the mean difference (−0.43–0.56) was also within the equivalence margin.

The KM MIC values showed a uniform and pronounced increase after the acquisition of AMK resistance associated with *rrs* mutations. Before the development of AMK resistance, the KM MIC values were in the range of 2–32 μg/mL, whereas all isolates exhibited KM MIC values >256 μg/mL thereafter, regardless of the testing medium (7H9 or CAMHB). Paired statistical analyses were not applicable because all post-acquisition MIC values exceeded the upper limit of the testing range. All isolates demonstrated a marked increase of at least +5–7 log₂ dilutions, indicating consistent and strong cross-resistance between AMK and KM.

### Antimicrobial susceptibility testing

The SM– and KM-resistant strains generated i*n vitro* were designated as 63S and 63 K, respectively. The MIC values of AMK, SM, and KM against 63S and 63 K cells were evaluated using the broth microdilution method (Table 3). In strain 63S, the SM MIC exceeded 256 µg/mL in both 7H9 and CAMHB, whereas the AMK and KM MIC values were in the range of 4–16 µg/mL. In strain 63 K, the MIC values of AMK and KM exceeded 256 μg/mL in both 7H9 and CAMHB broth, while that of SM was 16 μg/mL in both media.

**Table 3.** MIC results for AMK, SM, and KM in *Mycobacterium avium* isolates.

| Type of culture | Drug susceptibility test | Strain |  |  |
| --- | --- | --- | --- | --- |
|  |  | 63A<sup>a</sup> | 63S<sup>b</sup> | 63K<sup>c</sup> |
| CAMHB | AMK MIC (µg/mL) | 16 | 8 | >256 |
|  | SM MIC (µg/mL) | 16 | >256 | 16 |
|  | KM MIC (µg/mL) | 16 | 2 | >256 |
| 7H9 | AMK MIC (µg/mL) | 8 | 16 | >256 |
|  | SM MIC (µg/mL) | 4 | >256 | 16 |
|  | KM MIC (µg/mL) | 4 | 8 | >256 |

Abbreviations: AMK, amikacin; CAMHB, cation-adjusted Mueller-Hinton broth; KM, kanamycin; MIC, minimum inhibitory concentration; SM, streptomycin
<sup>a</sup> *Mycobacterium avium* strain isolated from a patient before treatment. <sup>b</sup> SM-resistant strain generated *in vitro* using 63A. <sup>c</sup> KM-resistant strain generated *in vitro* using 63A.

### Analysis of genetic mutations of high-level MIC of SM and KM isolates in a MAC clinical isolate

Table 4 summarizes the results of whole-genome SNPs and mutation analyses of strains 63 K and 63S, using 63A as the reference. Strain 63 K harbored two nucleotide substitutions: a C-to-G mutation within a gene encoding a hypothetical protein at position 350,310 and an A-to-G transition at position 1,488,929 corresponding to position 1408 of the *E. coli rrs* gene. Strain 63S exhibited a T-to-C substitution in a hypothetical protein gene at position 4,247,872 and a T-to-A mutation at positions 1,487,552, corresponding to position 20 of the *E. coli rrs* gene.

**Table 4.** SNPs and genetic mutations identified in 63K*^a^* and 63S*^b^* with 63A*^c^* as the reference genome.

|  | Position | Gene symbols | Annotation of genes | *E. coli rrs* position | Reference allele | Alternate allele |
| --- | --- | --- | --- | --- | --- | --- |
| 63K | 350310 | - | Hypothetical protein | - | C | G |
|  | 1488929 | *rrs* | 16S ribosomal RNA | 1408 | A | G |
| 63S | 4247872 | - | Hypothetical protein | - | T | C |
|  | 1487552 | *rrs* | 16S ribosomal RNA | 20 | T | A |

Abbreviations: SNP, single nucleotide polymorphism.
<sup>a</sup> KM-resistant strain generated *in vitro* using 63A. <sup>b</sup> SM-resistant strain generated *in vitro* using 63A. <sup>c</sup> *Mycobacterium avium* strain isolated from a patient before treatment.

### Streptomycin use in clinical practice

We encountered two cases in which SM remained clinically effective after the acquisition of AMK resistance associated with *rrs* mutations. In both cases, the SM MIC values at the time of confirmation of AMK resistance were ≤16 µg/mL. In one CAM-susceptible case, preoperative administration of SM for three months resulted in culture negativity and an improvement in radiological findings. After continued postoperative SM administration for an additional 3 months, culture negativity was maintained and the treatment was completed (Figure 1A, 1B). In another CAM-susceptible case, 4 months of SM administration were associated with decreased bacterial burden and improvement of pulmonary lesions on chest CT (Figure 2A, 2B).

**Fig 1.**
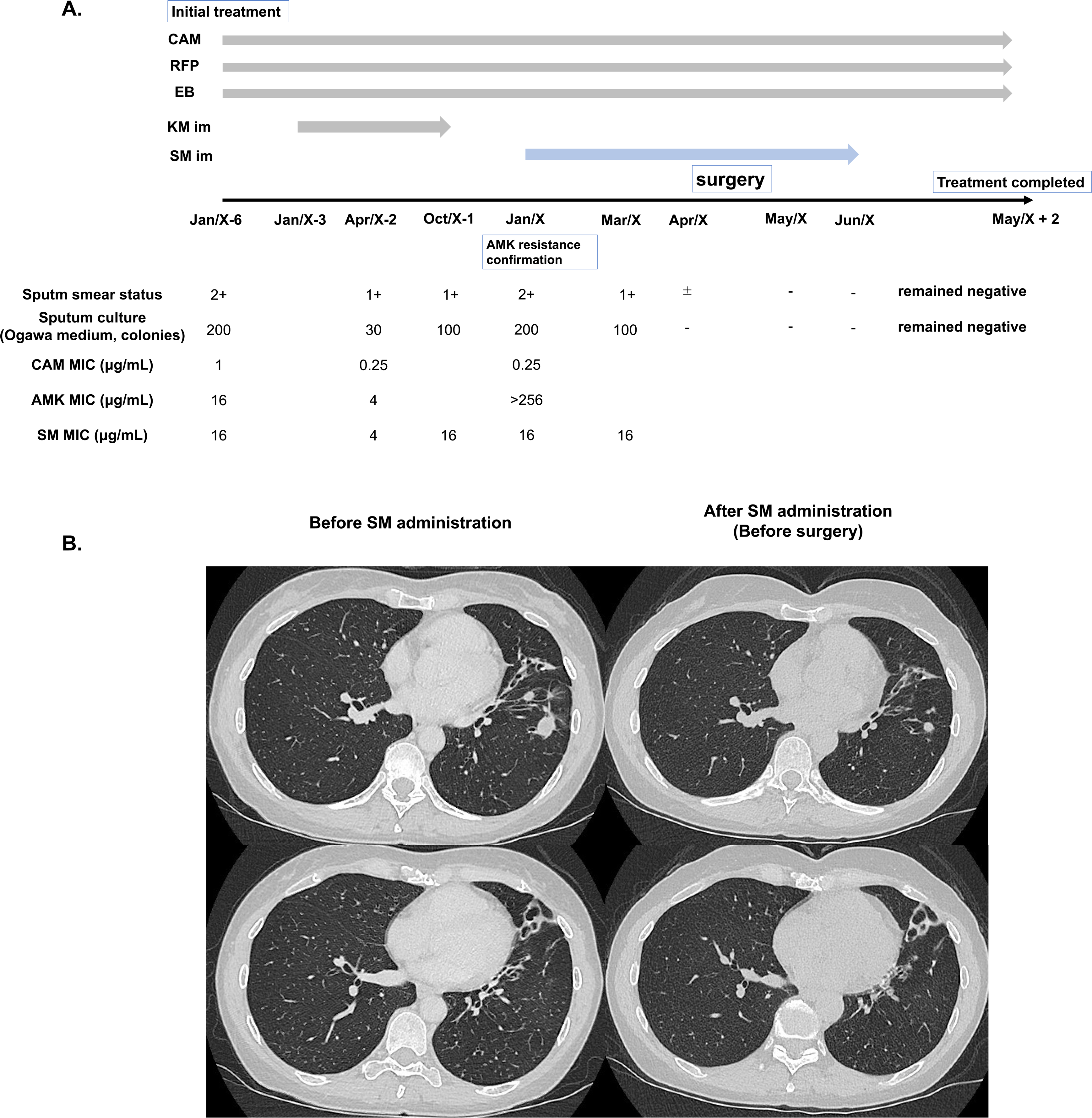
(A) The clinical course of a CAM-susceptible and AMK-resistant case in which negative conversion was achieved through pre– and post-operative administration of SM. (B) Chest computed tomography shows a reduction in small nodular and nodular opacities, as well as disseminated granular opacities, in the left lower lobe following SM administration; the left panel shows baseline findings before SM administration, and the right panel shows the pre-operative findings three months after SM administration.

**Fig 2.**
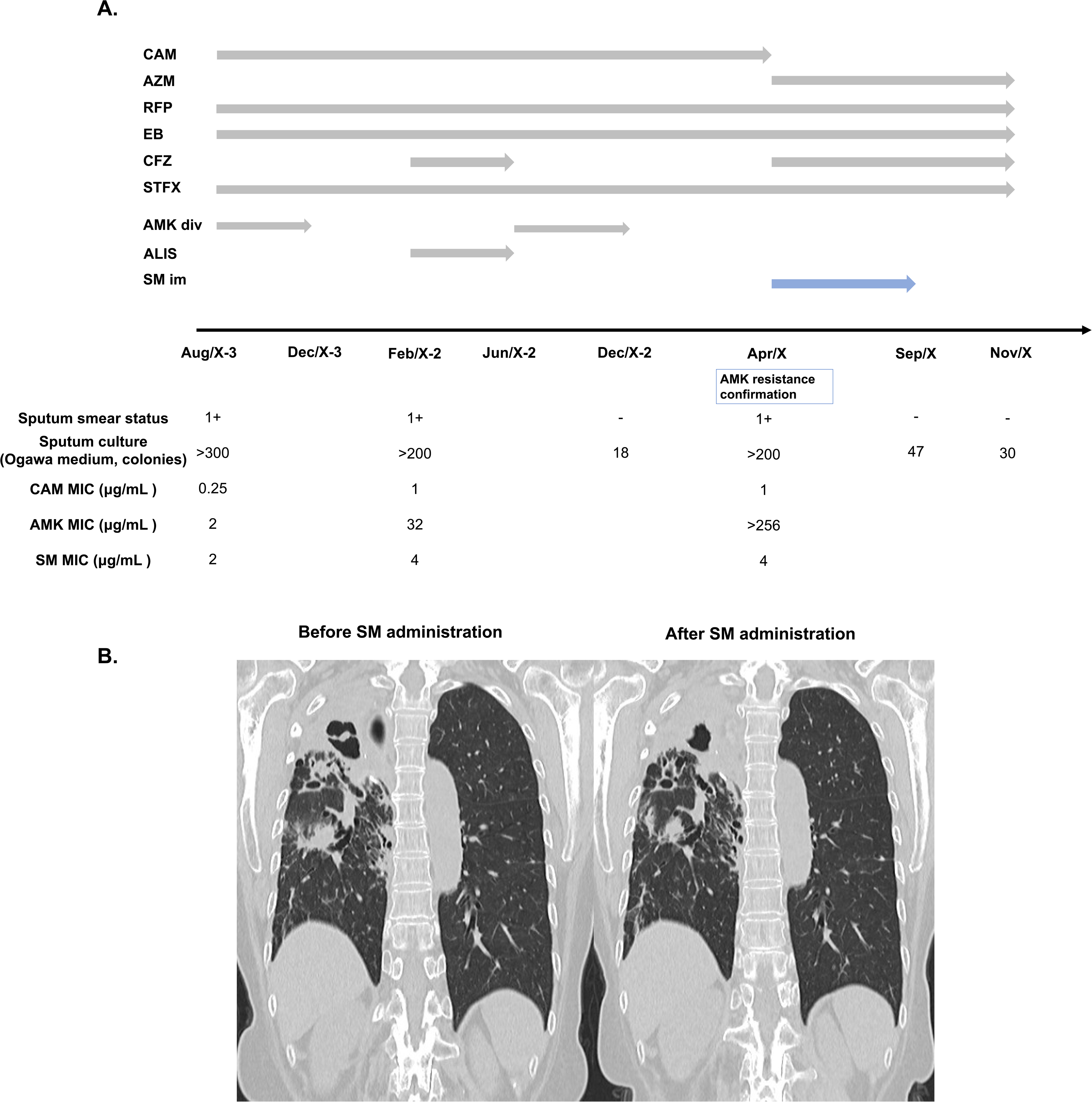
(A) The clinical course of a CAM-susceptible and AMK-resistant case in which the bacterial load decreased following SM administration. (B) Chest computed tomography shows a reduction of the cavitary lesion and infiltration in the right upper lobe following SM administration; the left panel shows baseline findings before SM administration, and the right panel shows findings four months after SM administration.

## DISCUSSION

To our knowledge, this is the first study to investigate the potential for cross-resistance among AMK, SM, and KM by assessing SM and KM MIC values in MAC strains before and after the acquisition of AMK resistance associated with *rrs* mutations, with the aim of identifying potential treatment options for AMK-resistant MACPD. The acquisition of AMK resistance did not result in a consistent and meaningful increase in SM MIC values. Furthermore, in isolates with high level SM MIC values generated *in vitro*, a mutation at position 20 of the *rrs* gene was identified; this site differed from the known AMK resistance–associated regions. These findings suggested a lack of cross-resistance between AMK and SM, which was strengthened in clinical cases treated with SM. In contrast, cross-resistance between AMK and KM was observed, with KM MIC values exceeding 256 µg/mL and shared *rrs* mutations associated with AMK resistance.

In this study, SM MIC values for CAMHB containing 5% OADC and Middlebrook 7H9 broth containing 10% OADC showed no significant changes after AMK resistance in 14 and 15 cases of AMK resistance associated with *rrs* mutations, respectively. Genome analysis of *in vitro*-created SM-resistant isolates showed a mutation at position 20 in the *rrs* gene, located at a site distinct from that in the previously reported AMK resistance-associated positions (7–12). Our data suggest that there is no cross-resistance between the two drugs in MAC, similar to the case in *M. tuberculosis* (20), suggesting that SM could remain a treatment alternative in AMK-resistant cases. Portillo-Gomez *et al*. did not identify genetic markers for SM resistance in MAC (25). Although we detected an *rrs* mutation at position 20 in an in vitro-generated SM-resistant isolate, this mutation may not represent all genetic mutation sites associated with SM resistance. Therefore, identification of SM-resistant isolates derived from clinical isolates is warranted for further analysis.

Regarding the efficacy of SM administration, a comparative study showed that adding SM during the first 3 months of macrolide-based oral therapy significantly improved sputum culture conversion in patients with MACPD compared with oral therapy alone (26). Recently, Kim *et al.* reported a similar conversion rate with SM– and AMK-containing regimens in patients with cavitary MACPD treated according to guideline-based treatment for at least 1 year (27). In our two AMK-resistant cases, both isolates had SM MIC values of ≤16 µg/mL, and the response to SM administration suggests potential therapeutic value. Although Kobashi *et al.* reported that the MIC values determined using Middlebrook 7H9 were not associated with clinical efficacy (28), the clinical significance of SM MIC values should be further clarified using a standardized method for MIC determination and an objective method of clinical evaluation. In this context, Fröberg *et al.* showed, using broth microdilution data from 12 laboratories, that the ECOFF for amikacin was 64 mg/L for MAC and that the current CLSI susceptible breakpoint for AMK (16 mg/L) divided the wild-type MIC distribution, highlighting that MIC interpretation remains affected by methodological and breakpoint-related limitations even for AMK (29). Therefore, before SM MIC values can be incorporated into routine therapeutic decision-making, susceptibility testing should be standardized. As the CLSI guidelines recommend CAMHB as the standardized medium for AMK MIC value determination, SM MIC testing should also be performed using CAMHB to support clinical decision-making.

Comparison of KM MIC values before and after the acquisition of AMK resistance showed that all isolates exhibited KM MIC values greater than 256 µg/mL in both Middlebrook 7H9 *broth* and CAMHB medium. Moreover, in a patient-derived isolate with a high KM MIC value *in vitro*, a mutation associated with AMK resistance was identified at the *rrs* gene associated with AMK resistance (7–12). These findings suggested that AMK and KM exhibited cross-resistance. The fact that two patients in this study received only KM as the AMG drug, in addition to standard oral therapy, further supports the possibility of cross-resistance between AMK and KM. Although KM was recommended along with SM for patients with refractory MACPD in 2012 in Japan (5), it is not currently recommended in the global treatment guidelines (4). Therefore, KM-induced AMK resistance is unlikely to be a major cause for concern.

Middlebrook 7H9 broth and CAMHB medium were used to determine the MIC values of each AMG drug. A previous study reported that the concordance rate of AMK MIC values between the two media types was only 18% (30). Here, the MIC values for each AMG drug, in the range of 0.25–256 µg/mL were evaluated using preserved isolates. When comparing the conditions before and after the confirmation of AMK resistance, the concordance rates for AMK and KM between 7H9 and CAMHB increased from 23.5% and 17.6%, respectively, to 100 %. In contrast, the concordance rate for SM remained low (29.4% before vs. 20.0% after). However, in one SM-resistant isolate, the SM MIC values were concordant (> 256μg/mL) between the two media. According to the CLSI guidelines, CAMHB is the recommended and standardized medium for AMK MIC value determination. Therefore, SM MIC testing should be performed using the CAMHB to support clinical decision-making.

This study had some limitations. This was a small single-center retrospective study. Our results should be confirmed in future multi-center studies. Second, this SM-resistant isolate was generated *in vitro*, and position 20 in the *rrs* mutation may not represent all the genetic mutation sites associated with SM resistance. Further studies on SM-resistant isolates derived from clinical specimens are required. Third, this study did not investigate the *in vitro* SM– and KM-resistant mutations in *Mycobacterium intracellulare.* In addition, the MIC data in 7H9 broth for the strains cultured before the patient was treated at Fukujuji Hospital were partially used for the analysis. Therefore, potential measurement variability could not be excluded despite the identical MIC testing methods.

In conclusion, AMK-resistant *M. avium-intracellular* isolates with *rrs* mutations do not confer resistance to SM. In addition, SM-resistant isolates generated *in vitro* did not exhibit AMK resistance, whereas KM-resistant strains did. These findings suggest that SM is a potential treatment alternative for selected patients with AMK-resistant MACPD. Furthermore, measurement of SM MIC values using CAMHB appears to be justified.

## Acknowledgements

We thank the staff of the clinical laboratory at the Fukujuji Hospital for managing the preserved strains.

## Author contributions

Tatsuya Kodama, Formal analysis, Data curation, Methodology, Writing–original draft

Kozo Morimoto, Conceptualization, Supervision, Writing–review & editing

Yoshiro Murase, Validation, Software

Akio Aono, Investigation, Methodology

Koji Furuuchi, Resources, Validation

Keiji Fujiwara, Resources, Validation

Masashi Ito, Resources, Validation

Takashi Ohe, Validation

Fumiya Watanabe, Methodology

Kinuyo Chikamatsu, Methodology

Shiomi Yoshida, Methodology

Yusuke Minato, Funding acquisition, Project administration

Yoshiaki Tanaka, Validation

Miyako Hiramatsu, Data curation

Yuji Shiraishi, Data curation, Writing–review & editing

Takashi Yoshiyama, Writing–review & editing

Satoshi Mitarai, Conceptualization, Supervision, Writing–review & editing

## Data availability statement

The sequencing reads have been deposited in the DDBJ Sequence Read Archive under accession number PRJDB40448, with run accessions DRR911509–DRR911511.

